# Early Prophylactic Hydrocortisone and Survival Without BPD in Extremely Preterm Infants

**DOI:** 10.1101/2025.07.20.25330756

**Authors:** Veronica Smedbäck, Lars J Björklund, Anders Flisberg, Jolanta Wróblewska, Olivier Baud, Erik Wejryd, Ulrika Ådén

**Affiliations:** Department of Biomedical and Clinical Sciences, Division of Children’s and Women’s health, Linköping University, Linköping, Sweden; Futurum, Ryhov County Hospital, Jönköping, Sweden; Pediatrics, Department of Clinical Sciences, Lund, Lund University, Lund, Sweden; Department of Pediatric Surgery and Neonatology, Skåne University Hospital, Lund and Malmö, Sweden; Institution of Clinical Sciences, Department of Pediatrics, Sahlgrenska Academy, Gothenburg, Sweden; The Queen Silvia Children’s Hospital, Department of Neonatology, Region Västra Götaland, Sahlgrenska University, Sweden; Department of Pediatrics, Umeå University, Umeå, Sweden; Obstetric, Perinatal, Paediatric and Life Course Epidemiology Team (OPPaLE), Center for Research in Epidemiology and StatisticS (CRESS), Alimentation Et L’Environnement (INRAe), Institut National Pour La Santé Et La Recherche Médicale (INSERM), French Institute for Health and Medical Research, Institut National de Recherche Pour L’Agriculture, l’Paris Cité University, Paris, France; Department of Neonatal Medicine, FHU Prem’Impact, Cochin-Port Royal Hospital, Assistance Publique-Hôpitaux de, Paris Cité University, 123 Bd de Port-Royal, 75014, Paris, France; Paris Cité University, Neuro-Diderot, INSERM, Paris, France; Department of Pediatrics, Vrinnevi hospital, Norrköping, Sweden; Department of Women’s and Children’s Health, Karolinska Institutet, Stockholm, Sweden; Department of Neonatal Medicine, Astrid Lindgrens Children’s Hospital. Karolinska University Hospital, Stockholm, Sweden; Department of Neonatal Medicine, H.K.H Crown Princess Victoria Children’s Hospital, Linköping University Hospital, Linköping, Sweden

## Abstract

**Importance:** In randomized trials, early prophylactic low-dose hydrocortisone improved survival without bronchopulmonary dysplasia (BPD) and had few adverse effects in extremely preterm infants. Large scale implementation data are needed to estimate effect size and safety.

**Objective:** To examine the association between early prophylactic hydrocortisone and survival without BPD at 36 weeks postmenstrual age in extremely preterm infants in Sweden after implementation, and to assess the safety of this treatment.

**Design:** A national historical cohort study with prospectively collected data.

**Setting:** Data was collected from the Swedish Neonatal Quality register from four Swedish regions where prophylactic hydrocortisone was implemented.

**Participants:** The study included 1140 infants born before 28 weeks gestation between 2018 and 2023. A total of 1106 infants met inclusion criteria. Infants were divided into exposed and non-exposed groups based on the intention-to-treat principle.

**Exposure:** Hydrocortisone 1 mg/kg/day for the first 7 days of life, followed by 0.5 mg/kg/day from days 8 to 10.

**Main outcomes and measures:** The primary outcome was survival without BPD at 36 weeks’ postmenstrual age. Logistic regression was used to present odds ratios (OR), both unadjusted and after adjustment for covariates.

**Results:** Among 1106 infants (median [IQR] gestational age, 25+6 [24+3-27+0] weeks; median [IQR] birth weight, 780 [610-964] g), 474 received prophylactic hydrocortisone and 632 did not. Survival without BPD occurred in 154 of 474 exposed infants (32.5%) and in 185 of 632 unexposed infants (29.3%). Adjusted OR was 1.62 (95% CI, 1.16–2.27). The reduction in BPD, rather than mortality, primarily drove this effect. The strongest association was observed in infants born at 24–25 weeks’ gestation. Late-onset bacterial infection was more common in the exposed group, but the difference was not significant after adjustment. No other severe neonatal morbidities differed significantly between the two groups.

**Conclusion and relevance:** Exposure to prophylactic hydrocortisone in extremely preterm infants was associated with increased survival without BPD, significant after adjustments. There was no significant increase in severe neonatal morbidities, except that late-onset bacterial infection was more common in the exposed group before adjustments.

**Key points:** *Question:* Does early prophylactic hydrocortisone improve survival without bronchopulmonary dysplasia in extremely preterm infants born in Sweden, and is it safe to use?

*Findings:* Using prospectively collected data from a national register, this study found that exposure to prophylactic hydrocortisone was associated with increased likelihood of survival without BPD. There was no significant increase in severe neonatal morbidities.

*Meaning:* This study, investigating real-world data, aligns with previous similar studies supporting the potential benefits and safety of early prophylactic hydrocortisone treatment.

## Introduction

Bronchopulmonary dysplasia (BPD) is a common chronic lung disease among infants born extremely preterm, defined as birth before 28 weeks’ gestation. Reported incidence rates vary widely, from 10% to 89% and, in Sweden, approximately 50-60% of all infants born extremely preterm are diagnosed with BPD at 36 weeks’ postmenstrual age (1-3). The multifactorial pathogenesis of BPD involves inflammation and is often associated with mechanical respiratory support. Extremely preterm infants are particularly vulnerable due to their immature adrenal function and low levels of endogenous cortisol. Combined with the postnatal stressors commonly encountered in the neonatal intensive care unit, this vulnerability and exposure increases the risk of developing BPD (4-6).

Numerous studies have indicated that corticosteroid replacement therapy may offer benefits for extremely preterm infants, by supporting hemodynamic stability, promoting lung development and reducing inflammation (7). Dexamethasone has historically often been used to or treat BPD, but early intervention is associated with a markedly increased risk of both short- and long-term adverse effects, including elevated risk for cerebral palsy (8-10).

Previous trials have documented positive effects of early prophylactic hydrocortisone on lung development and mortality, without causing neurodevelopmental impairment (11-14). The effect of prophylactic hydrocortisone to extremely preterm infants in clinical practice has previously been reported in three smaller single center studies (12, 15, 16). Thus, there is a need for real world data on the effectiveness of prophylactic hydrocortisone. Further, the treatment has been associated with adverse events, such as spontaneous intestinal perforation and late-onset sepsis. These complications may be related to interactions with other treatments or clinical practices such as concurrent use of indomethacin (9, 17, 18). Consequently, the safety aspects need to be further studied and evaluated in large-scale real-world settings.

## Objectives

This study aimed to analyze real-world data on the effect of early prophylactic hydrocortisone in extremely preterm infants in Sweden, where the treatment has been implemented in some regions since 2020. Our hypotheses were that prophylactic hydrocortisone would (1) increase survival without BPD at 36 weeks’ postmenstrual age and (2) that it would not be associated with increased rates of adverse effects or severe neonatal morbidities.

## Participants and methods

### Design/cohort

This study was a historical cohort study with prospectively collected data. Eligible infants were born before 28 weeks’ gestation and admitted to the neonatal intensive care unit on the first day of life at a hospital where prophylactic hydrocortisone had already been implemented (South, West, North, and South-east regions of Sweden). No exclusion criteria were applied for the study. Data from January 2018 to December 2023 were collected. The dates of prophylactic hydrocortisone implementation in each region were confirmed by local physicians. Infants born prior to the implementation date were used as controls. Data were obtained from the Swedish Neonatal Quality Register (SNQ), a national database that prospectively records daily clinical information on all infants admitted to neonatal care covering 98% (19) of all live births admitted for neonatal care in Sweden. Two sensitivity analyses were made, one with a broader national cohort including all infants born extremely preterm in Sweden, regardless of region, during 2018-2023 and the other with 1:1 matched propensity score groups. Both sensitivity analyses are presented in supplemental tables. The study was approved by the Swedish Ethical Review Authority (Dnr 2024-05118).

### Exposure

The exposure investigated in this study was prophylactic hydrocortisone, administered intravenously as hydrocortisone sodium succinate, 0.5 mg/kg twice daily from days 1 to 7 of life, followed by 0.5 mg/kg once daily for days 8 to 10, yielding a cumulative dose of 8.5 mg/kg. In cases where intravenous access was not available, hydrocortisone was administrated orally, typically toward the end of treatment. In three of the four regions, the treatment was given to all infants born before 28 weeks’ gestation. In one region (the North), the treatment was given to all infants born before 26 weeks’ gestation and to infants born at 26+0 to 27+6 weeks’ gestation if the mother had suspected chorioamnionitis. The validated dates of implementation were September 1, 2020 in the Western region; January 1, 2021 in the Southern and Northern regions; and April 1, 2022, in the Southeastern region. The two remaining regions in Sweden (East and Middle) have chosen not to adopt this treatment protocol.

### Outcomes

The primary outcome was survival without BPD at 36 weeks’ postmenstrual age. BPD was defined as the need for supplemental oxygen at 36 weeks’ postmenstrual age. Secondary outcomes included the total number of days in mechanical ventilator, age at completed weaning from respiratory support including airway support from mechanical ventilator, continuous positive airway pressure, high and low flow nasal cannula. Other secondary outcomes were patent ductus arteriosus (PDA) both medically and surgically treated, and the use of other systemic steroids. Safety outcomes included short term severe neonatal morbidities as defined by the register, including spontaneous intestinal perforation, necrotizing enterocolitis, late bacterial infection defined with positive bacterial cultures, pulmonary hemorrhage, insulin treatment, retinopathy of prematurity (ROP) diagnosis, treatment for ROP, intraventricular hemorrhage grade 3-4, and cystic periventricular leukomalacia.

### Statistical analysis

For descriptive analyses, Fisher exact tests were used for dichotomous variables, Chi-square tests for categorical variables, and t-test or Mann-Whitney U tests for continuous variables, depending on the distribution of the variable. Logistic regression was performed as the main analysis, both unadjusted and adjusted for relevant covariates, with odds ratios (OR) calculated along with 95% confidence intervals (CI) and p-values.

Sensitivity analyses were conducted using 1:1 propensity score (PS) matching based on all baseline characteristics, allowing balanced group comparisons without adjustments for covariates. A nearest neighbor matching algorithm with a caliper width of 0.2 of the standard deviation of the logit was applied with validation based on distribution scores between exposed and non-exposed before matching.

Continuous outcome variables were analyzed using linear regression, assuming normal distribution when applicable, or negative binomial regression depending on the distribution of the variables. Results from the negative binomial regression are presented as risk ratios (RR) with 95% CI and p-values. The number needed to treat to achieve survival without BPD was also calculated. All tests were two-tailed and conducted at a significance level of 0.05. Data were analyzed from July 2024 to February 2025 using SAS software version 9.4 (SAS Institute Inc., Cary, NC, USA). All analyses included only complete cases.

## Results

### Cohort

According to the Swedish Neonatal Quality Register, 2011 infants were born extremely preterm in Sweden between January 2018 and December 2023. Of these, 1140 were born in regions where prophylactic hydrocortisone was implemented (South, West, South-East, and North). A total of 1106 infants were eligible for the study. Inclusion criteria were admission to one of these neonatal intensive care units no later than on the first day of life. A total of 34 infants were excluded due to missing data in the register (n=34).

The infants were divided into groups of exposed and non-exposed to prophylactic hydrocortisone according to intention-to-treat principle, meaning that all infants born after the introduction of prophylaxis in the respective region were assumed to be treated and all infants born before that date were assumed not to be treated (Figure 1). The gestational age (GA) was significantly lower in the exposed group (median [IQR] GA 25+4 [24+2 to 27+0]) compared to the control group (median [IQR] 26+1 [24+4 to 27+1], p=0,02, table 1). No other significant baseline differences were observed. The median GA for the whole cohort was 25 weeks + 6 days (IQR 24+3 to 27+0) and median birth weight 780 grams (IQR 610-964 grams). All baseline characteristics are shown in Table 1. Baseline characteristics from the sensitivity analyses are detailed in eTable 1 for the propensity score matched groups and in eTable 2 for the larger national cohort.

**Table 1:** Baseline characteristics and group comparisons of exposed and non-exposed infants

| Baseline characteristics | Exposed group<br>(N=474) | Non-exposed group<br>(N=632) | p-value |
| --- | --- | --- | --- |
| GA, weeks (IQR) | 25+4 (24+2 - 27+0) | 26+1 (24+4 - 27+1) | 0.020 |
| GA 22-23 weeks, n (%) | 92 (19.4%) | 97 (15.3%) |  |
| GA 24-25 weeks, n (%) | 180 (38.0%) | 213 (33.7%) |  |
| GA 26-27 weeks, n (%) | 202 (42.6%) | 322 (50.9%) |  |
| Birth weight, g (IQR) | 755 (603 - 950) | 800 (610 - 968) | 0.23 |
| Male sex, n (%) | 257 (54.2%) | 346 (54.7%) | 0.90 |
| Prenatal steroids given, n (%) | 425/452 (94.0%) | 536/589 (91.0%) | 0.08 |
| Surfactant given, n (%) | 365 (77.0%) | 470 (74.4%) | 0.32 |
| Multiple births, n (%) | 99 (20.9%) | 159 (25.2%) | 0.18 |
| Intubation at birth, n (%) | 185/463 (40.0%) | 228/621 (36.7%) | 0.28 |
| Data are presented as median (interquartile range) or number (percentage). For test between two groups Fisher's exact test was used for dichotomous variables, Chi-square test for categorical variables, and Mann-Whitney U-test for continuous variables based on the variable distribution.<br>GA Gestational age. |  |  |  |

**Table 2:** The primary outcome and its components, with and without stratification

|  | Exposed group<br>N=474 | Control group<br>N=632 | OR<br>(95% CI) | p-value | aOR<br>(95% CI) | p-value |
| --- | --- | --- | --- | --- | --- | --- |
| Survival without BPD | 154 (32.5%) | 185 (29.3%) | 1.16<br>(0.90-1.50) | 0.25 | 1.62<br>(1.16 -2.27) | 0.0047 |
| Components of the primary outcome |  |  |  |  |  |  |
| BPD at 36 weeks PMA | 240 (50.6%) | 350 (55.4%) | 0.83<br>(0.65-1.05) | 0.12 | 0.68<br>(0.52-0.90) | 0.0064 |
| Death before 36 weeks PMA | 87 (18.4%) | 102 (16.1%) | 1.17<br>(0.85-1.60) | 0.33 | 1.12<br>(0.75-1.69) | 0.58 |
| Stratified by gestational age<sup>a</sup> |  |  |  |  |  |  |
| 22-23 weeks | 5 (5.4%) | 4 (4.1%) | 1.34<br>(0.35-5.14) | 0.67 | <sup>b</sup> | <sup>b</sup> |
| 24-25 weeks | 44 (24.4%) | 29 (13.6%) | 2.06<br>(1.23-3.47) | 0.0062 | <sup>b</sup> | <sup>b</sup> |
| 26-27 weeks | 105 (52.0%) | 152 (47.4%) | 1.20<br>(0.85-1.71) | 0.30 | 1.30<br>(0.85-1.99) | 0.23 |
| Stratified by birthweight deviation<sup>a</sup> |  |  |  |  |  |  |
| Small for gestational age | 24 (24.2%) | 30 (22.9%) | 1.08<br>(0.58-1.99) | 0.81 | 1.44 <sup>c</sup><br>(0.73-2.82) | 0.29 <sup>c</sup> |
| Normal birth weight | 129 (34.8%) | 152 (31.0%) | 1.19<br>(0.89-1.58) | 0.24 | 1.62<br>(1.1-2.35) | 0.011 |
| Stratified by chorioamnionitis<sup>a</sup> |  |  |  |  |  |  |
| Chorioamnionitis | 17 (34.0%) | 9 (20.0%) | 2.06<br>(0.81-5.25) | 0.13 | 7.78 <sup>c</sup><br>(1.72-35.14) | 0.0077 <sup>c</sup> |
| No chorioamnionitis | 137 (32.3%) | 176 (30.0%) | 1.11<br>(0.85-1.46) | 0.43 | 1.52<br>(1.08-2.15) | 0.017 |
| Stratified by sex<sup>a</sup> |  |  |  |  |  |  |
| Female | 79 (36.4%) | 95 (33.2%) | 1.15<br>(0.79-1.67) | 0.46 | 1.57<br>(0.96-2.57) | 0.07 |
| Male | 75 (29.2%) | 90 (26.0%) | 1.17<br>(0.82-1.68) | 0.39 | 1.65<br>(1.03-2.64) | 0.036 |
| Logistic regression was used unadjusted and adjusted for covariates.<br>Following covariates are used to adjust, except when covariate is a subgroup: sex, multiple births, birth weight with and without z-score, gestational age (weeks), Apgar score 10 min, intubation at birth, region of birth, prenatal steroids, and surfactant.<br><sup>a</sup> Not adjusted for region of birth.<br><sup>b</sup> Not enough number of events to make adjusted logistic regression analysis possible.<br><sup>c</sup> Only adjusted for gestational age.<br>*BPD* Bronchopulmonary dysplasia, *PMA* Postmenstrual age, *GA* Gestational age, *SGA* Small for gestational age, *OR* Odds ratio, *aOR* adjusted odds ratio, *CI* Confidence Interval. |  |  |  |  |  |  |

**Figure 1.**
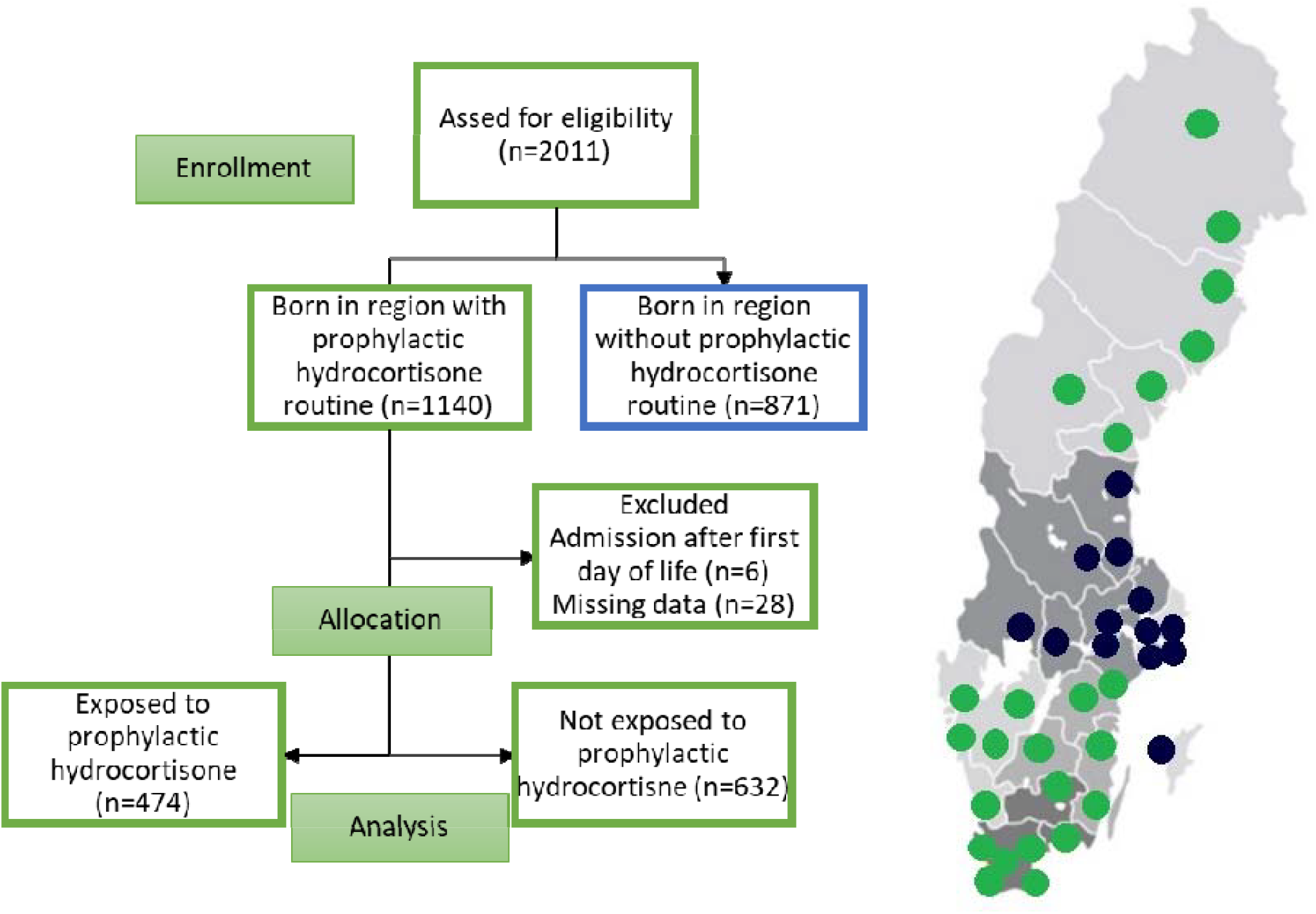
Flow chart of the cohort. The included map shows all neonatology units in Sweden, with green dots representing regions that have implemented early prophylactic hydrocortisone routine and blue dots representing regions without this routine.

### Main findings

#### Primary outcome

In the exposed group, 154 of 474 infants (32.5%) survived without BPD, compared with 185 of 632 infants (29.3%) in the non-exposed group. The unadjusted OR was 1.16 (95% CI, 0.90-1.50; p=0.25), and adjusted OR 1.62 (95% CI, 1.16-2.27; p=0.0047, Table 2). Covariates included in the adjusted logistic regression were GA (weeks), birth weight in gram and as z-score, sex, multiple births, prenatal steroids, Apgar at 10 min, intubation at birth, region of birth, and surfactant treatment. Sensitivity analyses were repeated with 1:1 propensity score matched groups which yielded a higher OR for survival without BPD between exposed and non-exposed groups (OR 1.40; 95% CI, 1.04-1.89, eTable 3).

**Table 3:** Secondary outcomes

| **Respiratory outcomes<sup>a</sup>** | **Exposed group**<br>**N=474** | **Control group**<br>**N=632** | **RR**<br>**(95% CI)** | **p-value** | **aRR**<br>**(95% CI)** | **p-value** |
| --- | --- | --- | --- | --- | --- | --- |
| Weaning from respiratory support, mean (95% CI) | 77 (73-83) | 73 (69-77) | 1.07<br>(0.98-1.16) | 0.13 | 1.03<br>(0.95-1.11) | 0.54 |
| Weaning from mechanical ventilation, mean (95% CI) | 33 (29-38) | 34 (30-38) | 0.98<br>(0.82-1.17) | 0.85 | 0.97<br>(0.81-1.16) | 0.73 |
| Mechanical ventilator days, mean (95% CI) | 12 (10-14) | 12 (11-14) | 0.96<br>(0.79-1.18) | 0.73 | 0.77<br>(0.63-0.93) | 0.0062 |
| **Other secondary outcomes<sup>b</sup>** | **Exposed group**<br>**N=474** | **Control group**<br>**N=632** | **OR**<br>**(95% CI)** | **p-value** | **aOR**<br>**(95% CI)** | **p-value** |
| Patent ductus arteriosus, medically treated, n (%) | 218 (46.0%) | 278 (44.0%) | 1.08<br>(0.85-1.38) | 0.51 | 0.89<br>(0.67-1.19) | 0.45 |
| Patent ductus arteriosus, surgically treated, n (%) | 26 (5.5%) | 39 (6.2%) | 0.88<br>(0.53-1.47) | 0.63 | 0.76^<br>(0.45-1.30) | 0.32<sup>c</sup> |
| Administration of systemic steroids other than prophylactic hydrocortisone, n (%) | 128 (27.0%) | 191 (30.2%) | 0.85<br>(0.66-1.11) | 0.24 | 0.77<br>(0.56-1.05) | 0.10 |

<sup>a</sup> Negative binomial regression was used unadjusted and adjusted for covariates. <sup>b</sup> Logistic regression was used unadjusted and adjusted for covariates. Following covariates are used to adjust for negative binomial- and logistic regression: sex, multiple births, birth weight with and without z-score, gestational age (weeks), Apgar score 10 min, intubation at birth, region of birth, prenatal steroids, and surfactant. <sup>c</sup> Only adjusted for gestational age. RR Relative risk, aRR Adjusted relative risk, CI Confidence Interval, OR Odds ratio, aOR adjusted odds ratio, CI Confidence Interval

In the larger national cohort including all six Swedish regions, the unadjusted OR for survival without BPD was 1.01 (95% CI 0.81-1.26) and the adjusted OR 1.58 (95% CI, 1.14–2.20, eTable 4).

**Table 4:**
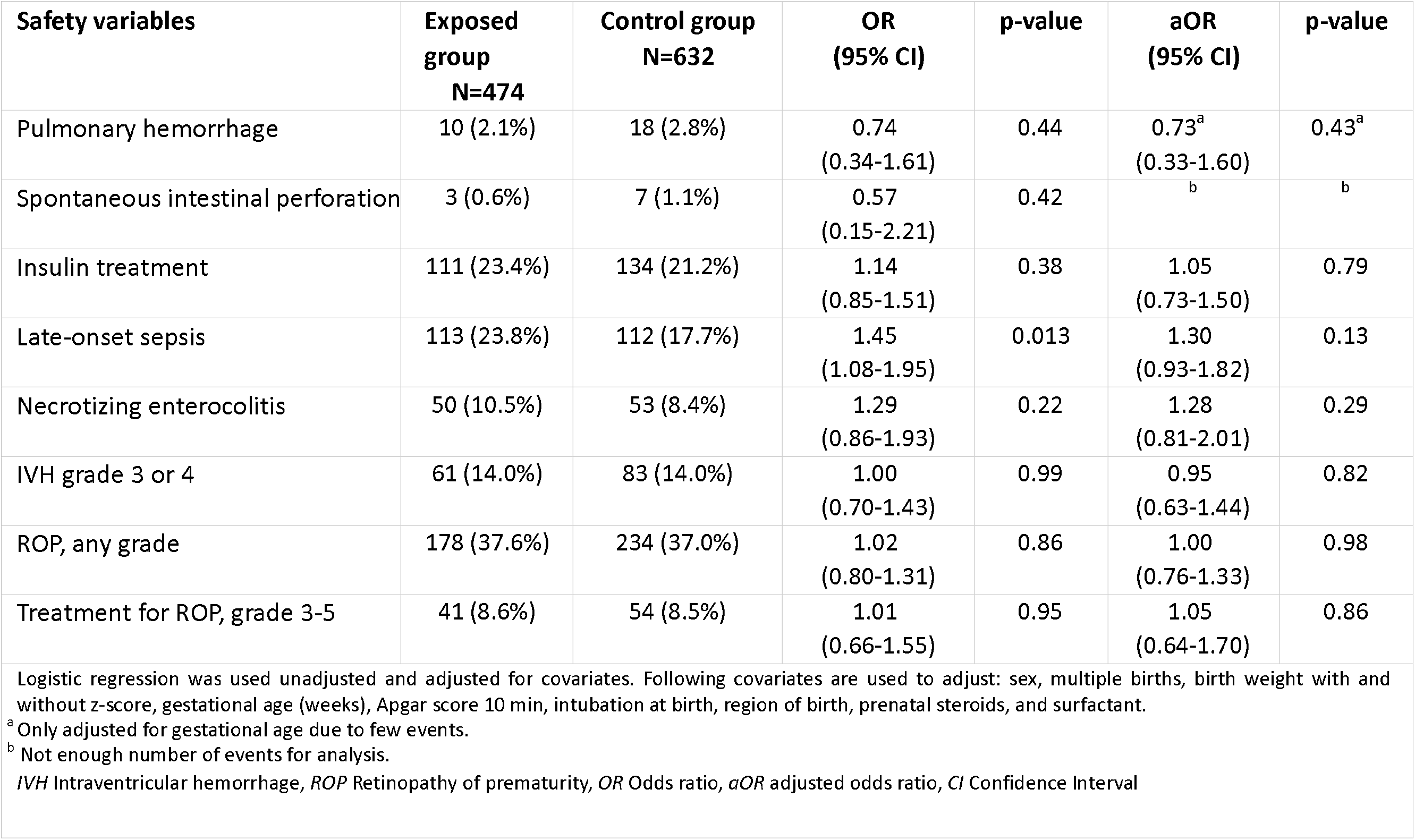
Outcome on safety variables.

The number needed to treat (NNT) to achieve one additional infant surviving without BPD was 32 based on unadjusted incidence rates, and 15 using the propensity score–matched groups.

When analyzing the components in the primary outcome (mortality before 36 weeks postmenstrual age, and BPD) we found that the OR for BPD between the two groups was OR 0.83 (95% CI, 0.65-1.05) and after adjustments OR 0.68 (95% CI, 0.52-0.90, Table 2). There was no significant difference between the exposed and non-exposed groups in mortality before 36 weeks postmenstrual age.

Stratified analysis by gestational age of primary outcome showed that the effect was most pronounced in infants born at 24–25 weeks gestation (OR, 2.06; 95% CI, 1.23–3.47, Table 2). Due to few eligible and included infants, adjusted regression could not be performed in this subgroup, but PS-matched analysis yielded an OR of 2.51 (95% CI, 1.33–4.75). Additional subgroup analyses by birth weight, presence of chorioamnionitis, and sex are shown in Table 2. In the national cohort, similar reductions in OR for BPD were observed (adjusted OR, 0.69; 95% CI, 0.53-0.91), as well as increased survival without BPD in infants born at 24–25 weeks gestation (OR, 1.56; 95% CI, 1.03-2.35, eTable 4).

#### Secondary outcomes

There were no significant differences between the exposed and unexposed groups in the incidence of PDA, neither overall nor among infants in need of medical or surgical treatment for PDA. This finding remained consistent across unadjusted and adjusted logistic regression, as well as in propensity score–matched analyses (Table 3).

Infants in the exposed group had significantly fewer days on mechanical ventilation compared to non-exposed infants after adjustments (adjusted relative risk [aRR], 0.77; 95% CI, 0.63–0.93, Table 3). No significant differences were observed in the unadjusted analysis or in other respiratory variables, including time to weaning from respiratory support. The use of systemic corticosteroids other than prophylactic hydrocortisone did not differ significantly between the two groups (aOR, 0.77; 95% CI, 0.56–1.05, Table 3).

#### Safety analyses

There were no significant differences between exposed and non-exposed groups in the incidence of spontaneous intestinal perforation, necrotizing enterocolitis, insulin treatment, pulmonary hemorrhage, intraventricular hemorrhage, treatment for ROP, or cystic periventricular leukomalacia (Table 4).

Late-onset bacterial infection occurred more frequently in the group exposed to prophylactic hydrocortisone compared with the non-exposed (23.8% vs 17.7%; unadjusted OR 1.45, 95% CI, 1.08-1.95), but this difference was no longer statistically significant after adjustment for covariates (aOR 1.30, 95% CI, 0.93-1.82, Table 4). No significant differences were observed in propensity score– matched analyses regarding safety variables although late-onset bacterial infection was close to statistically significant (OR 1.39, 95% CI, 0.99-1.94, eTable 5). In the larger national cohort, both late-onset bacterial infection and insulin treatment were more common in the exposed group; however, these associations were not statistically significant after covariate adjustment or propensity score matching (eTable 6).

## Discussion

In this national historic cohort study, based on prospectively collected register data, exposure to early prophylactic hydrocortisone in extremely preterm infants was associated with significantly increased survival without BPD after adjustment for covariates. This benefit was not accompanied by an increased risk of severe neonatal morbidities.

In the unadjusted analysis, the incidence of late-onset bacterial infection was more common in the exposed group compared to the non-exposed group, consistent with findings from the PREMILOC trial (11). After adjusting for covariates and using propensity score–matched analyses, the difference was no longer statistically significant, although a trend persisted. Shah et al. (12) also investigated the safety of prophylactic hydrocortisone in extremely preterm infants and similarly found no significant association with rates of positive bacterial cultures and prophylactic hydrocortisone.

Neither our study nor that of Shah et al. identified a significant association between prophylactic hydrocortisone and bowel perforation, despite prior reports suggesting such a link in certain trials (14, 17, 18, 20). This findings in our Swedish cohort might be explained by the limited use of indomethacin, a treatment for PDA that has been implicated in increasing the risk of bowel perforation (9, 21).

To minimize confounding, we employed two distinct control strategies. The first used historical controls from the same regions, reducing bias from well-known regional differences in clinical practice. The second included a national cohort from all six regions, allowing for comparisons with contemporary infants unexposed to prophylactic hydrocortisone, reducing the potential impact of improvements in clinical practices over time. In both strategies, we applied propensity score matching to further reduce confounding as a sensitivity analysis. All outcomes were significant after adjusting for covariates in the cohort, the overall larger cohort and propensity score-matched groups. The persistence of similar outcomes despite these adjustments strengthens our conclusions, even though statistical significance was only achieved post-adjustments.

Previous randomized controlled trials have demonstrated reduced mortality with early prophylactic hydrocortisone, with mixed effects on BPD (8). However, both our findings and those of Shah et al. suggest that the main driver of increased survival without BPD is a reduction in BPD incidence, rather than improved survival. One speculation is that this difference may reflect overall improvements in neonatal care and lower baseline mortality in recent years, making it more difficult to demonstrate a mortality benefit (2, 22, 23).

The timing of prophylactic hydrocortisone may also play a crucial role. Previous studies have shown that the fetal adrenal gland continues to mature until full term, and that extreme preterm birth is associated with cortisol insufficiency. Therefore, the effectiveness of hydrocortisone replacement for cortisol insufficiency may depend on the timing of exposure (24, 25). We observed a stronger association between prophylactic hydrocortisone and increased survival without BPD in infants born at 24–25 weeks of gestation. This more pronounced improvement in the smallest infants support the hypothesis that early low-dose prophylactic hydrocortisone serves as a replacement therapy for the cortisol deficiency caused by immature adrenals.

It is possible that a similar benefit may exist for infants born at 22–23 weeks’ gestation; however, our study may have been underpowered to detect a statistically significant effect in this subgroup due to few events resulting in wider confidence intervals. Additionally, this study examined subgroups for the primary outcome; small for GA, chorioamnionitis, and male sex. Our hypothesis was that exposure to prophylactic hydrocortisone would have a greater effect in these subgroups, but few events may also have limited the statistical power for these analyses.

While previous studies have reported that prophylactic hydrocortisone may reduce the need for PDA treatment (9, 11, 12), we did not observe such an association. A possible explanation for this may be the discrepancy in treatment practices for PDA in different countries, which could lead to population level differences in treatment thresholds or preferences and thereby limit cross-study comparability (26).

The current study lacks validation of the BPD diagnosis, which was defined as the need for supplemental oxygen at 36 weeks postmenstrual age. Other definitions of BPD have been used in studies such as the PREMILOC trial and the study by Shah et al., making direct comparisons challenging. Despite this, our findings are broadly consistent with the PREMILOC trial and other observational studies.

This study has several strengths. As a registry-based study, it benefits from a large, nationwide population sample derived from a high-quality registry with near-complete data capture (19).

Furthermore, the use of intention-to-treat approach minimizes selection bias, including exclusion of infants who may have died early before treatment initiation. Finally, representation from all participating regions and ongoing author collaboration helped reduce potential regional biases There are limitations to this study. The lack of a validated BPD diagnosis reduces the reliability of our findings and prevents categorization of BPD according to more contemporary definitions, including severity. Conversely, BPD definition based on need for supplemental oxygen at 36 weeks postmenstrual age is a rather pragmatic definition frequently used in clinical practice. Further, the severity of BPD may be particularly relevant, given that recent study linked dexamethasone exposure to improved survival without cerebral palsy in infants with severe BPD (27). The effect of prophylactic hydrocortisone exposure on severe BPD and its impact on survival without cerebral palsy remains unclear. Additionally, the current study relied on registry data collected by clinicians, limiting our ability to independently verify data quality or content. Nevertheless, data reporting to the registry has been ongoing for over 20 years with set standards and daily submission protocols (19).

Finally, the study emphasizes the need for further research, ideally through prospective studies, to identify whether specific subgroups of extremely preterm infants may derive greater benefit or face harm from early prophylactic hydrocortisone therapy. Additionally, follow-up studies are particularly important to assess unknown neurodevelopmental effects and is at the moment ongoing by the authors.

## Conclusion

This national cohort study provides evidence that exposure to early prophylactic hydrocortisone in extremely preterm infants is associated with increased survival without BPD after adjustments, without any registered short-term side effects. Long-term follow-up and further prospective studies are warranted to assess potential risks and benefits, particularly regarding neurodevelopmental outcomes and identification of high-risk subgroups.

## Data Availability

All data produced in the present study are available upon reasonable request to the authors

## Acknowledgments

Mrs Veronica Smedbäck, Drs Erik Wejryd and Ulrika Ådén had full access to all the data in the study and take responsibility for the integrity of the data and the accuracy of the data analysis. The study was supported by the Cocozza Foundation and research grant from ALF at region Östergötland. We thank Aldina Pivodic, Christopher Backström and Hussnain Khalid for their contribution to statistical analysis and interpretation of data. This work was presented orally and by poster at European Academy of Paediatric Societies in Vienna 2024 and Pediatric Academic Societies in Honolulu 2025.

