## Supplementary material for "Early Prophylactic Hydrocortisone and Survival Without BPD in Extremely Preterm Infants": Suplemental tables

### Supplementary tables

**eTable 1: Baseline characteristics with propensity score matched groups analyses presented**

| **Baseline characteristics** | **Exposed group (N=474)** | **Non-exposed group (N=632)** | **p-value** | **PS 1:1 p-value (N=419/419)** |
| --- | --- | --- | --- | --- |
| GA, weeks (IQR) | 25+4 (24+2 – 27+0) | 26+1 (24+4 – 27+1) | **0.020** | 0.49 |
| GA 22-23 weeks, n (%) | 92 (19.4%) | 97 (15.3%) |  |  |
| GA 24-25 weeks, n (%) | 180 (38.0%) | 213 (33.7%) |  |  |
| GA 26-27 weeks, n (%) | 202 (42,6%) | 322 (50,9%) |  |  |
| Birth weight, g (IQR) | 755 (603-950) | 800 (610-986) | 0.23 | 0.63 |
| Male sex, n (%) | 257 (54.2%) | 346 (54.7%) | 0.90 | 0.94 |
| Prenatal steroids given, n (%) | 425 (94.0%) | 536 (91.0%) | 0.08 | 0.32 |
| Surfactant given, n (%) | 365 (77.0%) | 470 (74.4%) | 0.32 | 0.52 |
| Multiple births, n (%) | 99 (20,9%) | 159 (25,2%) | 0.18 | 0.56 |
| Intubation at birth, n (%) | 185 (40.0%) | 228 (36.7%) | 0.28 | 0.89 |
| Data are presented as median (interquartile range) or number (percentage). For test between two groups Fisher’s exact test was used for dichotomous variables, Chi-square test for categorical variables, and Mann-Whitney U-test for continuous variables based on the variable distribution.  Following covariates are used to create propensity score matched groups: gestational age (weeks), birth weight, Apgar score 10 min, sex, multiple births, intubation at birth, prenatal steroids, chorioamnionitis and surfactant.  *GA* Gestational age. | | | | |

**eTable 2: Baseline characteristics with cohort of infants from all Swedish regions born extremely preterm 2018-2023**

| **Baseline characteristics** | **Exposed group (N=474)** | **Non-exposed group (N=1510)** | **p-value** | **PS 1:1 p-value (N=425/425)** |
| --- | --- | --- | --- | --- |
| GA, weeks (IQR) | 25+4 (24+2 – 27+0) | 25+6 (24+3 – 27+0) | **0.048** | 0.17 |
| GA 22-23 weeks, n (%) | 92 (19.4%) |  |  |  |
| GA 24-25 weeks, n (%) | 180 (38.0%) |  |  |  |
| GA 26-27 weeks, n (%) | 202 (42.6%) |  |  |  |
| Birth weight, g (IQR) | 755 (603-950) | 780 (610-964) | 0.33 | 0.29 |
| Male sex, n (%) | 257 (54.2%) | 828 (54.8%) | 0.90 | 0.37 |
| Prenatal steroids given, n (%) | 425 (94.0%) | 1286 (90.6%) | **0.026** | 0.26 |
| Surfactant given, n (%) | 365 (77.0%) | 1129 (74.8%) | 0.36 | 0.81 |
| Multiple births, n (%) | 99 (20.9%) | 319 (21.1%) | 0.92 | 0.21 |
| Intubation at birth, n (%) | 185 (40.0%) | 626 (42.9%) | 0.28 | 0.89 |
| Data are presented as median (interquartile range) or number (percentage). For test between two groups Fisher’s exact test was used for dichotomous variables, Chi-square test for categorical variables, and Mann-Whitney U-test for continuous variables based on the variable distribution.  Following covariates are used to create propensity score matched groups: gestational age (weeks), birth weight, Apgar score 10 min, sex, multiple births, intubation at birth, prenatal steroids, chorioamnionitis and surfactant.  *GA* Gestational age. | | | | |

**eTable 3: The primary outcome and its components, with and without stratification and with propensity score matched groups analyses presented**

| **Primary Outcome** | **Exposed group, N=474** | **Control group, N=632** | **OR (95% CI)** | **p-value** | **aOR (95% CI)** | **p-value** | **PS 1:1 OR (95% CI) (N=419/419)** | **p-value** |
| --- | --- | --- | --- | --- | --- | --- | --- | --- |
| Survival without BPD | 154 (32.5%) | 185 (29.3%) | 1.16 (0.90-1.50) | 0.25 | **1.62 (1.16 -2.27)** | **0.0047** | **1.40 (1.04-1.89)** | **0.028** |
| BPD at 36 weeks PMA | 240 (50.6%) | 350 (55.4%) | 0.83 (0.65-1.05) | 0.12 | **0.68 (0.52-0.90)** | **0.0064** | **0.76 (0.58-0.99)** | **0.044** |
| Death before 36 weeks PMA | 87 (18.4%) | 102 (16.1%) | 1.17 (0.85-1.60) | 0.33 | 1.12 (0.75-1.69) | 0.58 | 1.07 (0.74-1.54) | 0.71 |
| **Stratified by gestational age ^a^** | | | | | | | | |
| 22-23 GA | 5 (5.4%) | 4 (4.1%) | 1.34 (0.35-5.14) | 0.67 | **^b^** | **^b^** | 1.49 (0.34-6.46) | 0.60 |
| 24-25 GA | 44 (24.4%) | 29 (13.6%) | **2.06 (1.23-3.47)** | **0.0062** | **^b^** | **^b^** | **2.51 (1.33-4.75)** | **0.0047** |
| 26-27 GA | 105 (52.0%) | 152 (47.4%) | 1.20 (0.85-1.71) | 0.30 | 1.30 (0.85-1.99) | 0.23 | 1.37 (0.91-2.05) | 0.13 |
| **Stratified by birthweight deviation ^a^** | | | | | | | | |
| SGA | 24 (24.2%) | 30 (22.9%) | 1.08 (0.58-1.99) | 0.81 | 1.44 **^c^** (0.73-2.82) | 0.29 **^c^** | 1.37 (0.68-2.78) | 0.38 |
| Normal birthweight | 129 (34.8%) | 152 (31.0%) | 1.19 (0.89-1.58) | 0.24 | **1.62 (1.1-2.35)** | **0.011** | **1.42 (1.02-1.98)** | **0.039** |
| **Stratified by chorioamnionitis ^a^** | | | | | | | | |
| Chorioamnionitis | 17 (34.0%) | 9 (20.0%) | 2.06 (0.81-5.25) | 0.13 | **7.78 ^c^ (1.72-35.14)** | **0.0077 ^c^** | 2.10 (0.76-5.81) | 0.15 |
| No chorioamnionitis | 137 (32.3%) | 176 (30.0%) | 1.11 (0.85-1.46) | 0.43 | **1.52 (1.08-2.15)** | **0.017** | 1.34 (0.98-1.84) | 0.06 |
| **Stratified by gender ^a^** | | | | | | | | |
| Female | 79 (36.4%) | 95 (33.2%) | 1.15 (0.79-1.67) | 0.46 | 1.57 (0.96-2.57) | 0.07 | 1.35 (0.88-2.07) | 0.17 |
| Male | 75 (29.2%) | 90 (26.0%) | 1.17 (0.82-1.68) | 0.39 | **1.65 (1.03-2.64)** | **0.036** | 1.35 (0.88-2.07) | 0.17 |
| Logistic regression was used unadjusted and adjusted for covariates for Overall and unadjusted for PS 1:1 matched groups. Following covariates are used to adjust: sex, multiple births, birth weight with and without z-score, gestational age (weeks), Apgar score 10 min, intubation at birth, region of birth, prenatal steroids, and surfactant.  Following covariates are used to create propensity score matched groups: gestational age (weeks), birth weight, Apgar score 10 min, sex, multiple births, intubation at birth, prenatal steroids, chorioamnionitis and surfactant.  **^a^** Not adjusted for region of birth.  **^b^** Not enough number of events to make adjusted logistic regression analysis possible.  **^c^** Only adjusted for gestational age  *BPD* Bronchopulmonary dysplasia, *PMA* Postmenstrual age, *GA* Gestational age, *SGA* Small for gestational age, *OR* Odds ratio, *CI* Confidence Interval. | | | | | | | | |

**eTable 4: The primary outcome and its components, with and without stratification with cohort on all infants born extremely preterm 2018-2023**

| **Primary Outcome** | **Exposed group, n=474** | **Control group, n=1510** | **OR (95% CI)** | **p-value** | **aOR (95% CI)** | **p-value** | **PS 1:1 OR (95% CI) (N=425/425)** | **p-value** |
| --- | --- | --- | --- | --- | --- | --- | --- | --- |
| Survival without BPD | 154 (32.5%) | 487 (32.3%) | 1.01 (0.81-1.26) | 0.92 | **1.58 (1.14 -2.20)** | **0.0065** | **1.36 (1.01-1.83)** | **0.042** |
| BPD at 36 weeks PMA | 240 (50.6%) | 762 (50.5%) | 1.01 (0.82-1.24) | 0.95 | **0.69 (0.53-0.91)** | **0.0090** | 0.77 (0.59-1.01) | 0.06 |
| Death before 36 weeks PMA | 87 (18.4%) | 275 (18.2%) | 1.01 (0.77-1.32) | 0.94 | 1.14 (0.77-1.70) | 0.52 | 1.09 (0.76-1.56) | 0.65 |
| **Stratified by gestational age ^a^** | | | | | | | | |
| 22-23 GA | 5 (5.4%) | 16 (6.1%) | 0.89 (0.32-2.49) | 0.82 | **^b^** | **^b^** | 1.99 (0.37-10.59) | 0.42 |
| 24-25 GA | 44 (24.4%) | 85 (17.2%) | **1.56 (1.03-2.35)** | **0.035** | **^b^** | **^b^** | **2.52 (1.33-4.76)** | **0.0044** |
| 26-27 GA | 105 (52.0%) | 386 (51.3%) | 1.03 (0.75-1.40) | 0.86 | 1.30 (0.85-2.00) | 0.23 | 1.42 (0.95-2.11) | 0.09 |
| **Stratified by birthweight deviation ^a^** | | | | | | | | |
| SGA | 24 (24.2%) | 73 (23.7%) | 1.03 (0.61-1.75) | 0.91 | 1.26 **^c^** (0.71-2.23) | 0.44 **^c^** | 1.45 (0.70-3.01) | 0.32 |
| Normal birthweight | 129 (34.8%) | 406 (34.4%) | 1.02 (0.80-1.30) | 0.89 | **1.57 (1.09-2.27)** | **0.015** | 1.39 (0.99-1.90) | 0.06 |
| **Stratified by chorioamnionitis ^a^** | | | | | | | | |
| Chorioamnionitis | 17 (34.0%) | 36 (27.7%) | 1.35 (0.67-2.71) | 0.41 | 1.43 **^c^** (0.59-3.44) | 0.43 **^c^** | 1.93 (0.68-5.44) | 0.21 |
| No chorioamnionitis | 137 (32.3%) | 451 (32.7%) | 0.98 (0.78-1.24) | 0.89 | **1.50 (1.07-2.12)** | **0.020** | 1.32 (0.97-1.80) | 0.08 |
| **Stratified by gender ^a^** | | | | | | | | |
| Female | 79 (36.4%) | 245 (35.9%) | 1.02 (0.74-1.40) | 0.90 | 1.49 (0.91-2.44) | 0.12 | 1.45 (0.94-2.25) | 0.09 |
| Male | 75 (29.2%) | 242 (29.2%) | 1.00 (0.73-1.36) | 0.99 | **1.61 (1.02-2.53)** | **0.039** | 1.45 (0.94-2.25) | 0.09 |
| Table 2: Outcome on primary outcome with and without stratifications. Logistic regression was used unadjusted and adjusted for covariates for Overall and unadjusted for PS 1:1 matched groups. Following covariates are used to adjust: sex, multiple births, birth weight with and without z-score, gestational age (weeks), Apgar score 10 min, intubation at birth, region of birth, prenatal steroids, and surfactant.  Following covariates are used to create propensity score matched groups: gestational age (weeks), birth weight, Apgar score 10 min, sex, multiple births, intubation at birth, prenatal steroids, chorioamnionitis and surfactant.  **^a^** Not adjusted for region of birth.  **^b^** Not enough number of events to make adjusted logistic regression analysis possible.  **^c^** Only adjusted for gestational age  *BPD* Bronchopulmonary dysplasia, *PMA* Postmenstrual age, *GA* Gestational age, *SGA* Small for gestational age, *OR* Odds ratio, *CI* Confidence Interval. | | | | | | | | |

**eTable 5: Outcome on safety variables with propensity score matched groups analyses presented**

| **Safety variables** | **Exposed group N=474** | **Control group N=632** | **OR (95% CI)** | **p-value** | **aOR (95% CI)** | **p-value** | **PS 1:1 OR (95%CI)** | **p-value** |
| --- | --- | --- | --- | --- | --- | --- | --- | --- |
| Pulmonary hemorrhage | 10 (2.1%) | 18 (2.8%) | 0.74 (0.34-1.61) | 0.44 | 0.73 **^a^** (0.33-1.60) | 0.43 **^a^** | 0.61 (0.25-1.48) | 0.27 |
| Spontaneous intestinal perforation | 3 (0.6%) | 7 (1.1%) | 0.57 (0.15-2.21) | 0.42 | **^b^** | **^b^** | 0.50 (0.12-2.00) | 0.32 |
| Insulin treatment | 111 (23.4%) | 134 (21.2%) | 1.14 (0.85-1.51) | 0.38 | 1.05 (0.73-1.50) | 0.79 | 1.14 (0.83-1.58) | 0.41 |
| Late-onset sepsis | 113 (23.8%) | 112 (17.7%) | **1.45 (1.08-1.95)** | **0.013** | 1.30 (0.93-1.82) | 0.13 | 1.39 (0.99-1.94) | 0.06 |
| Necrotizing enterocolitis | 50 (10.5%) | 53 (8.4%) | 1.29 (0.86-1.93) | 0.22 | 1.28 (0.81-2.01) | 0.29 | 1.41 (0.88-2.25) | 0.16 |
| IVH grade 3 or 4 | 61 (14.0%) | 83 (14.0%) | 1.00 (0.70-1.43) | 0.99 | 0.95 (0.63-1.44) | 0.82 | 1.11 (0.73-1.68) | 0.62 |
| ROP, any grade | 178 (37.6%) | 234 (37.0%) | 1.02 (0.80-1.31) | 0.86 | 1.00 (0.76-1.33) | 0.98 | 1.15 (0.87-1.52) | 0.32 |
| Treatment for ROP, grade 3-5 | 41 (8.6%) | 54 (8.5%) | 1.01 (0.66-1.55) | 0.95 | 1.05 (0.64-1.70) | 0.86 | 1.16 (0.72-1.88) | 0.54 |
| Logistic regression was used unadjusted and adjusted for covariates for Overall and unadjusted for PS 1:1 matched groups. Following covariates are used to adjust: sex, multiple births, birth weight with and without z-score, gestational age (weeks), Apgar score 10 min, intubation at birth, region of birth, prenatal steroids, and surfactant.  Following covariates are used to create propensity score matched groups: gestational age (weeks), birth weight, Apgar score 10 min, sex, multiple births, intubation at birth, prenatal steroids, chorioamnionitis and surfactant.  **^a^** Only adjusted for gestational age due to few events.  **^b^** Not enough number of events to make adjusted logistic regression analysis possible.  *IVH* Intraventricular hemorrhage, *ROP* Retinopathy of prematurity, *OR* Odds ratio, *aOR* adjusted odds ratio, *CI* Confidence Interval, *PS* Propensity score | | | | | | | | |

**eTable 6: Outcome on safety variables with cohort of infants from all Swedish regions born extremely preterm 2018-2023**

| **Safety variables** | **Exposed group N=474** | **Control group N=1510** | **OR (95% CI)** | **p-value** | **aOR (95% CI)** | **p-value** | **PS 1:1 OR (95%CI)** | **p-value** |
| --- | --- | --- | --- | --- | --- | --- | --- | --- |
| Pulmonary hemorrhage | 10 (2.1%) | 42 (2.8%) | 0.75 (0.38-1.51) | 0.43 | 0.75 **^a^** (0.37-1.51) | 0.43 **^a^** | 0.80 (0.31-2.04) | 0.63 |
| Spontaneous intestinal perforation | 3 (0.6%) | 23 (1.5%) | 0.41 (0.12-1.38) | 0.15 | 0.41 **^a^** (0.12-1.36) | 0.14 **^a^** | 0.50 (0.12-2.00) | 0.32 |
| Insulin treatment | 111 (23.4%) | 197 (13.0%) | **2.04 (1.57-2.64)** | **<0.001** | 1.06 (0.75-1.51) | 0.74 | 1.11 (0.81-1.53) | 0.52 |
| Late-onset sepsis | 113 (23.8%) | 271 (17.9%) | **1.43 (1.12-1.84)** | **0.0047** | 1.34 (0.96-1.88) | 0.08 | 1.31 (0.94-1.83) | 0.11 |
| Necrotizing enterocolitis | 50 (10.5%) | 160 (10.6%) | 0.99 (0.71-1.39) | 0.98 | 1.30 (0.83-2.04) | 0.25 | 1.40 (0.88-2.22) | 0.16 |
| IVH grade 3 or 4 | 61 (14.0%) | 188 (13.4%) | 1.05 (0.77-1.39) | 0.77 | 0.97 (0.64-1.45) | 0.86 | 1.01 (0.67-1.52) | 0.95 |
| ROP, any grade | 178 (37.6%) | 565 (37.4%) | 1.01 (0.81-1.24) | 0.96 | 1.03 (0.78-1.36) | 0.83 | 1.20 (0.91-1.58) | 0.20 |
| Treatment for ROP, grade 3-5 | 41 (8.6%) | 114 (7.5%) | 1.16 (0.80-1.68) | 0.44 | 1.06 (0.65-1.72) | 0.82 | 1.43 (0.86-2.37) | 0.16 |
| Logistic regression was used unadjusted and adjusted for covariates for Overall and unadjusted for PS 1:1 matched groups. Following covariates are used to adjust: sex, multiple births, birth weight with and without z-score, gestational age (weeks), Apgar score 10 min, intubation at birth, region of birth, prenatal steroids, and surfactant.  Following covariates are used to create propensity score matched groups: gestational age (weeks), birth weight, Apgar score 10 min, sex, multiple births, intubation at birth, prenatal steroids, chorioamnionitis and surfactant.  **^a^** Only adjusted for gestational age due to few events.  *IVH* Intraventricular hemorrhage, *ROP* Retinopathy of prematurity, *OR* Odds ratio, *aOR* adjusted odds ratio, *CI* Confidence Interval, *PS* Propensity score | | | | | | | | |
